# Mobile Cabin Hospital compulsory quarantine for mild patients was an alternative method to combat COVID-19: the Chinese experience

**DOI:** 10.1101/2020.07.26.20162206

**Authors:** Working Group on Fighting the COVID-19 epidemic in WuHan Hanyang Mobile Cabin Hospital, China, Hongru Li, Jiaping Lin, Hongmei Lian, Kang Chen, Yongtao Lyu, Yusheng Chen, Lili Ren, Li Zheng, Zhisheng Lin, Xueying Yu, Zihan Chen, Hend Al-Wathnani, Christopher Rensing, Xin Qian, Xinghai Yang

**Affiliations:** Shengli Clinical Medical College of Fujian Medical University, NO 134, Dongjie Street, Gulou District, Fuzhou City, 350001,P.R.China; Department of Pulmonary and Critical Care Medicine, Fujian Provincial Hospital, Fuzhou 350001, China; Department of Emergency Center, Fujian Provincial Hospital, Fuzhou 350001, China; China(Fujian) National Emergency Medical Rescue Team; Hebei Women and Children hospital; China(Sichuan)National Emergency Medical Rescue Team; Shandong Provincial Third Hospital; NHC Key Laboratory of Systems Biology of Pathogens and Christophe Mérieux Laboratory, Institute of Pathogen Biology, Chinese Academy of Medical Sciences & Peking Union Medical College, Beijing 100730, P. R. China; Institute of Environmental Microbiology, College of Resources and Environment, Fujian Agriculture & Forestry University, Fuzhou 350002, China; Institute of Urban Environment, Chinese Academy of Sciences, 1799 Jimei Road, Xiamen 361021, China; Department of Botany and Microbiology, King Saud University, Riyadh, Saudi Arabia

**Keywords:** Infectious diseases, novel coronavirus, coronavirus disease 2019 (COVID-19), mobile cabin hospitals, prevention and control measures

## Abstract

**Background:** Since the end of 2019 to the present day, the outbreak of the coronavirus disease 2019 (COVID-19) has had an immense impact on China and on other countries worldwide. This outbreak represents a serious threat to the lives and health of people all around the world. The epidemic first broke out in Wuhan, where the Chinese government was unable to prevent the spread of the disease by implementing home quarantine measures. Mobile cabin hospitals were used to relieve pressure on hospitals due to the need for beds while also isolating the sources of the infection through a centralized quarantine and treatment of mild cases.

**Method:** This paper reviewed and summarized the treatment of patients with mild illness and symptoms during the period from the construction to the closing of the Hanyang Mobile Cabin Hospital in Wuhan, China, and presented the operational elements and possible improvements of running this hospital.

**Results:** Mobile cabin hospitals helped China to curb the epidemic in only 2 incubation periods in 28 days.The basic conditions required for a normal operation of mobile cabin hospitals included the selection of the environment, medical staff to patient ratio, organizational structure, management model, admission criteria, treatment approaches, discharge process, livelihood guarantee, security, and other safeguarding measures. All of these components were performed carefully in Wuhan Hanyang Mobile Cabin Hospital, without medical staff being infected.

**Conclusion:** The mobile cabin hospital compulsory quarantine for mild patients was an alternative method to combat COVID-19. It is hoped that the presented work in this manuscript can serve as a reference for the emergency prevention and control measures for global epidemic outbreaks.

## Introduction

At the end of 2019, there was an outbreak of coronavirus disease 2019 (COVID-19) epidemic in Wuhan, China. Due to its long incubation period, atypical symptoms, multiple organ dysfunction due to multiple organ involvement, and insufficient medical resources in the early stages, the mortality rate of COVID-19 was estimated at 6%-10% ^[1-5].^ Furthermore, COVID-19 displayed an unprecedented rate of transmission, with a basic reproduction number (R0) of about 2-3 ^[6-8]^, hence the number of cases rose sharply from 41 cases in December 30, to 14,557 in February 3, including 14,411 cases in China and 146 cases in 22 countries outside of China ^[9]^. This led to a series of problems, such as a shortage of hospital beds, intense pressure on medical resources due to the exponential growth in the number of cases, and the difficulties faced by designated hospitals due to an overflow of patients. In the early stages of the epidemic outbreak, the Chinese government adopted home quarantine measures for patients with mild symptoms. However, outbreaks in familial clusters became one of the major routes of transmission in the development of the epidemic at the later stages ^[3]^ and the previously adopted home quarantine approach was shown to be unable to effectively control the epidemic. The disease continued spreading within many families, and mild cases could not be effectively contained, which then in turn became important sources of infection. This vicious cycle sustained the spread of the disease, resulting in an ongoing exponential growth of new and suspected cases (Figure 1). Secondly, due to insufficient medical resources, a large number of patients were unable to receive effective treatment, which caused more patients to become critically ill and increased the mortality rate ^[10-12]^. The situation became increasingly dire, and new strategies were urgently needed to cope with the situation.

**Figure 1:**
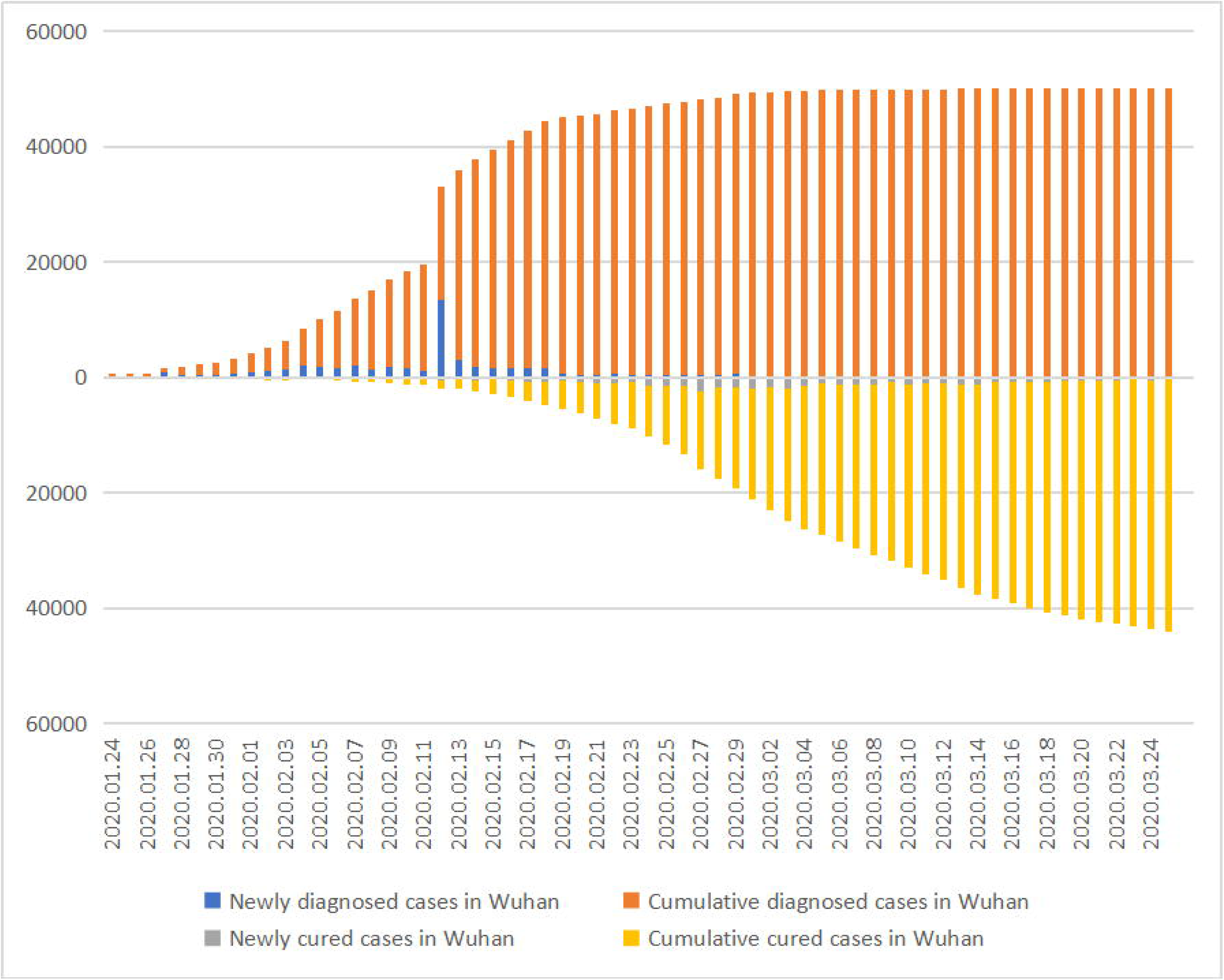
Demographics of newly diagnosed and cured cases, cumulative diagnoses and cured cases over time in Wuhan. Since the opening of the mobile cabin hospital on Feb 5th, the number of newly diagnosed cases in Wuhan (capital of Hubei province), the center of the epidemic, gradually decreased. This decrease could also be observed in Hubei Province, and the rest of China. Moreover, 13436 cases were clinical diagnosed on Feb 12th due to emergency strategy in this special time according to diagnosis and treatment protocol of novel coronaviral pneumonia (trial version5) in China. The number of newly diagnosed cases dropped to below 1000 and newly cured cases increased in Wuhan and the two lines crossed on Feb 20th, suggesting an inflection point of the epidemic. Moreover, the rate of cumulative diagnosed cases has gradually slowed down, and leveled off from then on. As the number of cured cases increased, the number of existing confirmed cases decreased significantly, suggesting that the epidemic had been effectively curbed. ---Data from the Chinese National Health Commission

To further control the sources of infection and prevent the deterioration of patients’ conditions, On February 3, China’s central government decided to implement the Mobile Cabin Hospital policy for confirmed cases with mild disease symptoms for concentrating quarantine and treatment, in addition to the policy that suspected cases and patients with fever underwent centralized quarantine (one patient, one room) through the requisition of hotels, guest houses, idle factories etc., before testing, diagnosis, and treatment. The so-called mobile cabin hospitals were wartime field hospitals that were built by converting large sports centers and exhibition centers for the purposes of receiving mild cases. This approach, on the one hand, enabled the unified centralized quarantine, treatment, and management of patients with mild disease symptoms, thus reducing transmission. On the other hand, it meant that designated hospitals were used for the treatment of severe cases, which enabled the concentration of resources and treatment, thereby increasing the recovery rate and decreasing the case fatality rate.

From February 3 to March 10, a total of 14 mobile cabin hospitals were set up and used in China, providing a total of 12,365 beds ^[16]^. The mobile cabin hospitals had been in operation for 35 days as of the writing of this paper, and a total of 11,309 patients had received treatment(Table1). The construction of mobile cabin hospitals had had a significant effect on reducing transmission and slowing down the growth rate of new cases (see Figure1). Since they had operated for an incubation period of 14 days, the newly diagnosis cases dropped significantly, for two incubation periods the number of new cases had gradually fallen to single digits. On the other hand, with an increase in the number of recovered patients, and the current number of confirmed cases having decreased substantially (Figure1), the trend of a rapid spreading the epidemic disease was curbed quickly as a result. Therefore, mobile cabin hospitals had played a crucial role in China’s success in the fight against the epidemic.

The specific names of the mobile cabin hospitals, the number of beds, and the number of patients admitted were listed in Table 1.

The authors came from a working team in Wuhan HanYang Moblie Cabin Hospital.The team composed of members from a Local hospital named Hubei Women and Children Hospital which was responsible for the management of the temporary hospital and 2 National Emergency Medical Rescue Teams from Fujian Province and Sichuan Province and two medical teams from Shandong and Sichuan. this paper contains a description of the authors as a first-hand witness account of the main conditions required for the normal operation of mobile cabin hospitals.

## Method

This paper reviewed and summarized the treatment of patients with mild illness during the period from the construction to the closing of the Hanyang Mobile Cabin Hospital in Wuhan, China, and proposed the operation elements and possible improvements of running this hospital.

## Results

### The structures and performance of HanYang Moblie Cabin Hospital

#### I. Location selection and ward layout description

Wuhan Hanyang Mobile Cabin Hospital was transformed by Hall B1 in Wuhan International Expo Center [Figure 2a]. Hanyang Mobile Cabin Hospital(MCH) was located in Hall B1, while the headquarter was set in Hall A1, which was in the upstream of Hall B1 and considered as clean zone. There were some measures taken before it began to run in order to prevent the cross infection of medical staff and patients and ensure the smooth operation(see Figure 2). First, the route toward the MCH for entering and leaving of Medical staff (blue arc line with arrow) was designed to be separated from that for patients (red arc line with arrow)(Figure 2a). Secondly, three zones and two passage ways were set up in MCH to facilitate the management of hospital infection control. (1)Two passage ways included one for doctors (? and another for patients ? which were completely separated. (2) The three zones included contaminated zone (red), buffer zone (yellow), and clean zone (outside the MCH, not shown). The contaminated zone contained preview triage area and the ward. In the buffer zone where doctors’ passageway ? was located, there was an entry channel (a) to don PPE (1 changing room, 1 buffer rooms) and an exit channel (b) to dof PPE. The exit channel also had four rooms: 1 the first buffer room, 2 the first doffing room, 3 the second doffing room, and 4 the second buffer room(Figure 2b). While the area outside the Hall B1, for example, Hall A1 was designated as the clean zone, and used as working area. Thirdly, In the middle of buffer area, a temporary CT reading area was set to facilitate the clinician to read patient’s chest imaging(Figure 2b)(see supplemetal material1). Lastly, in front of the MCH, in the patients’ pathway, there were two mobile CT scanners for patients’ imaging test with a waiting area in a tent. On the left, there was a simple clinical laboratory for blood testing that had been transformed from a convenience store(see supplemental material2).

**Figure 2.**
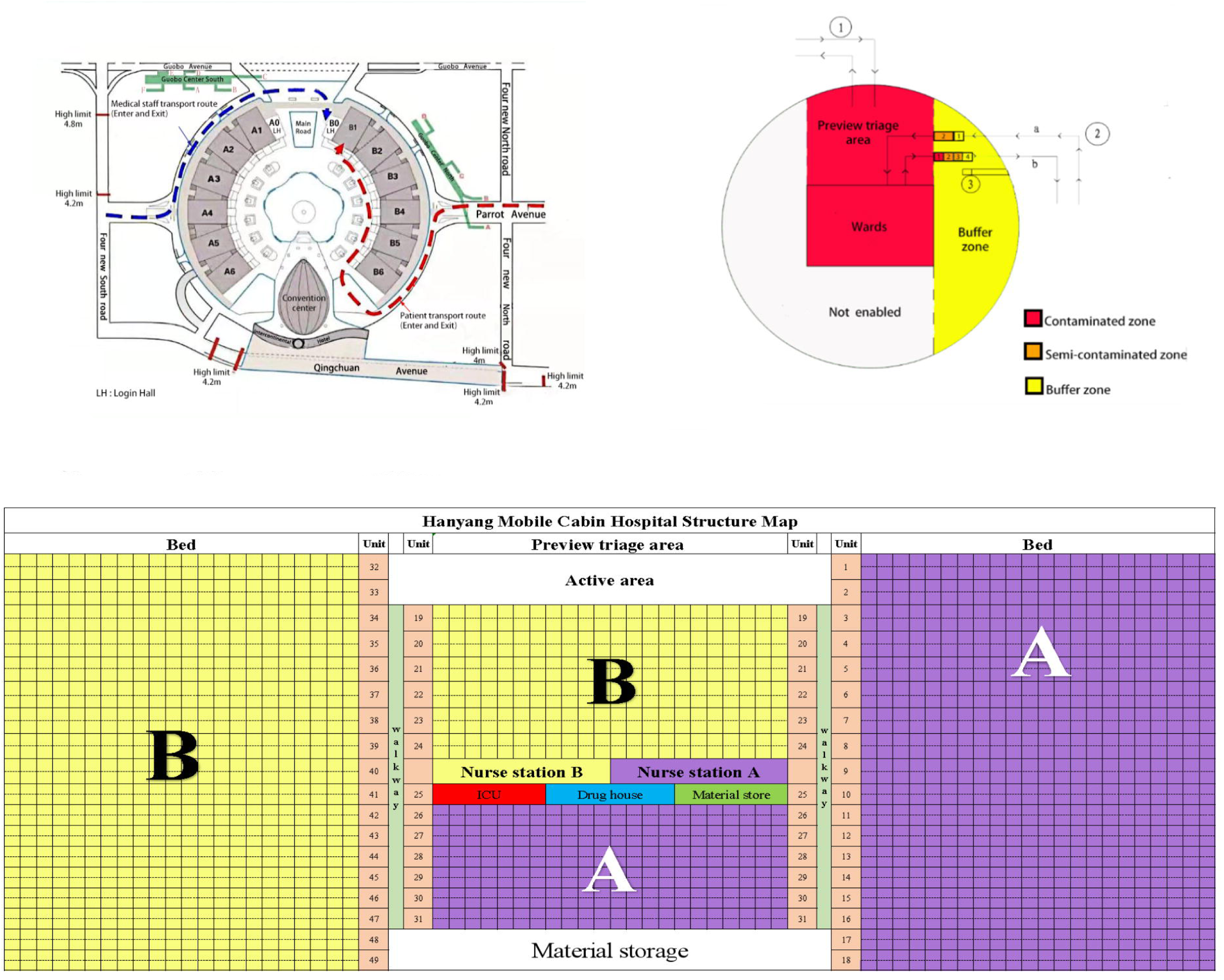
Location and structure of Wuhan Hanyang Mobile Cabin Hospital. Figure 2a: Map displaying the structure of Wuhan International Expo Center and Location of Hanyang Mobile Cabin Hospital. Hanyang Mobile Cabin Hospital was located in Hall B1, while the headquarter was in Hall A1.The transport route for entering and leaving of Medical staff (blue arc line with arrow) was designed to be separated from patients (red arc line with arrow) to prevent coinfection. Figure 2b: Graphic display of the design of Hanyang Mobile Cabin Hospital(Hall B1): three zones and two passage ways were set up to facilitate the management of hospital infection control. Two passage ways included one for doctors □ and another for patients □ which were completely separated. The three zones included contaminated zone (red), buffer zone (yellow), and clean zone (not shown). The contaminated zone contained preview triage area and the ward. In the buffer zone where doctors’ passageway □ was located, there was an entry channel (a) to don PPE (1 changing room, 1 buffer rooms) and an exit channel (b) to dof PPE. The exit channel also had four rooms: 1 the first buffer room, 2 the first doffing room, 3 the second doffing room, and 4 the second buffer room. While the area outside the Hall B1, for example, Hall A1 was designated as the clean zone, and used as working area. □ Passage way for patients;□ Passage way for doctors a: entry channel b: exit channel;□ CT reading zone .PPE: personal protective equipment. Figure 2c: The internal structure of the ward: there were 48 units which consisted of 18-22 beds in each unit, making up a total of 930 beds in the ward. The general ward was divided into two parts named A and B, managed by Shandong medical team and Sichuan medical team respectively. The Nurse Station was located in the central part, on the opposite side facing the Nurse Station, there were the intensive care unit (ICU), the Drug House, and the material storage room. There were reading room and activity area in the front of the ward, while at the back it was equipped with the storage area.

**Figure 3:**
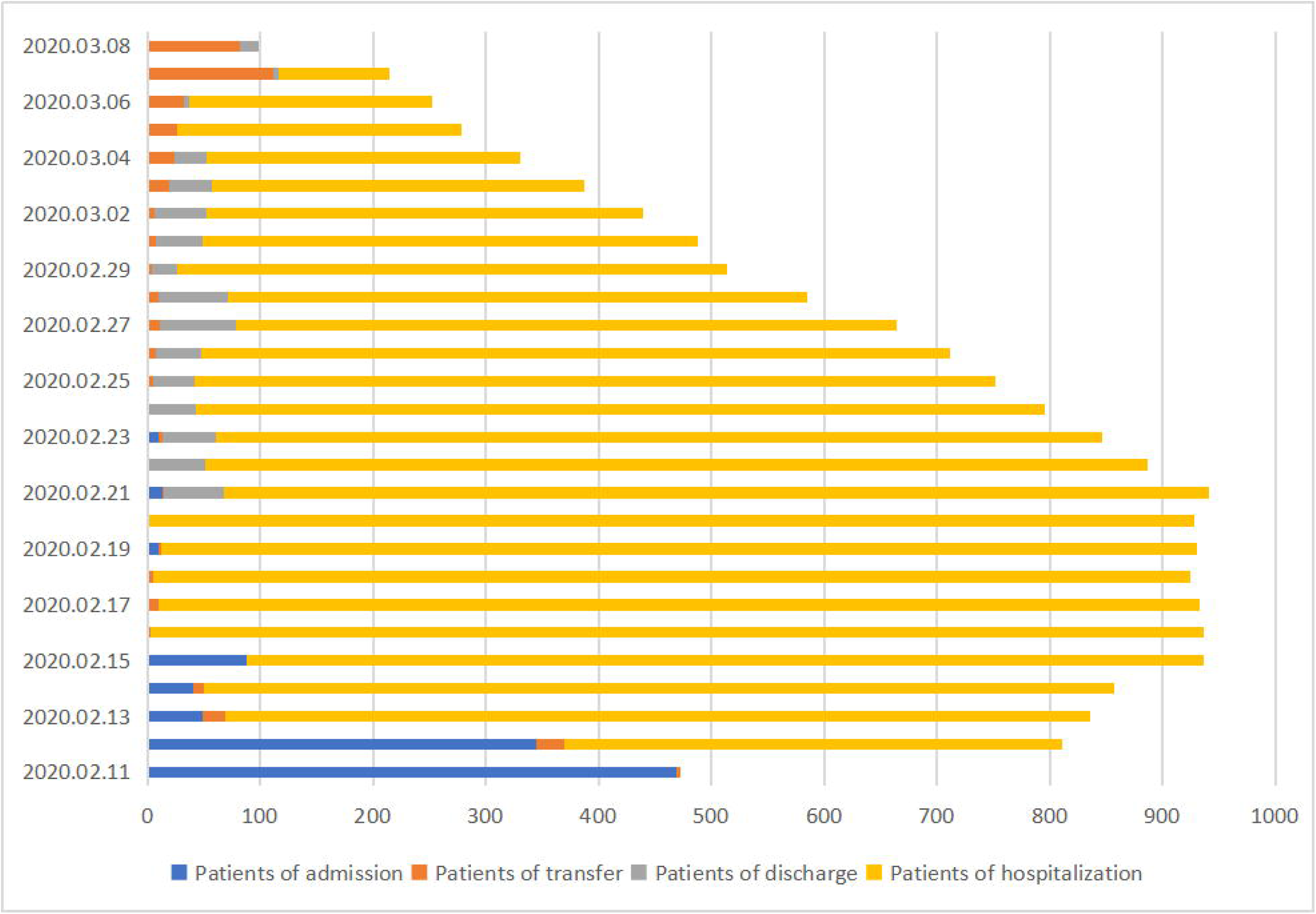
Trend of Patients undergoing hospitalization, admission, transfer and discharge in Hanyang Mobile Cabin Hospital. As was shown in Figure 3, a large number of patients were admitted at an early time on Feb 11, causing the mobile cabin hospital to be filled rapidly in just four days. There were about 50 patients discharged every day about 10 days later when the patients gradually recovered. In total, about 40% of patients were transferred to designated hospitals for treatment due to the severity of Covid-19 infections or other underlying basic diseases. There were nearly 200 critically ill patients transferred to designated hospitals on March 8, when the cabin hospital was closed because the epidemic in Wuhan had been curbed.

#### II. Ward layout and requirements for medical staff

There were 48 units which consisted of 18-22 beds in each unit, making up a total of 930 beds in the ward. The general ward was divided into two parts named A and B, managed by Shandong medical team and Sichuan medical team respectively. The Nurse Station was located in the central part, on the opposite side facing the Nurse Station, there were the intensive care unit (ICU), the Drug House, and the material storage room. There were reading room and activity area in the front of the ward, while at the back it was equipped with the storage area (Figure 2C)(see nhsupplemetal material3).

Bed spacing was at least 1-1.5m ^[17]^, Hanyang Mobile Cabin Hospital had two medical teams called Shangdong and Sichuan Medical Team, each of which managed 465 beds. Each team had about 300 medical staff including 100 physicians and 200 nurses who were divided into several groups. Therefore, the ratio of physicians to the nurses to patients was approximately 1:2:5. Every team had several groups for shifting works in MCH. The medical group leader (second-line physician) in each group managed the entire patients, and each front-line physician managed about 40 patients, hence at least 12 physicians were needed for each shift in each medical group. Additionally, special hospital infection teams were also established, scheduled for four shifts per day and four stations per shift, which included contaminated zones, semi-contaminated zones, buffer zones and cleaning stations (where material distribution, cleaning of googles and other work was performed). These teams were in charge of supervising the proper wearing and removal of personal protective equipment (PPE) for all types of workers to ensure zero infection.

#### III. The admission and operation of Hanyang Mobile Cabin Hospital

According to the “Workbook of mobile cabin Hospitals (Third Edition)” released by the Bureau of Hospital Administration, Department of Medical Administration, National Health Commission of China, the patients admitted to mobile cabin hospitals should be mild cases with mild clinical symptoms and no manifestations of pneumonia on imaging findings; and common cases with fever and respiratory symptoms, some manifestations of pneumonia on imaging findings, had self-care ability and were independently ambulatory. Patients should be free of severe chronic diseases, including hypertension, diabetes, coronary heart disease, malignant tumors, structural lung disease, pulmonary heart disease, and immunosuppression. Patients should not have a history of mental illness. Their resting-state finger pulse oximetry SpO2 should be above 93% and respiratory rate should be <30 breaths/min. Special instructions are given for other conditions ^[16]^.

In Hanyang MCH with A total of 930 beds, 1,028 patients were treated, 598 patients were cured, and 430 patients were transferred to designated hospitals before the MCH was closed.

As was shown in Figure3A, large number of patients were admitted at an early time on Feb 11, due to the outbreak being at an early stage, the mobile cabin hospital filled up rapidly in just four days.

Novel coronavirus nucleic acid testing was performed in the mobile cabin hospital starting on Feb 17th based on the diagnosis and treatment protocol of novel coronaviral pneumonia (trial version 7) in China, when the patients gradually recovered. Novel coronavirus nucleic acid testing in HanYang MCH was performed by KingMed Diagnostics Company. In Figure 4a, the results indicated there were about 30-120 patients who met discharge criteria with two negative nucleic acid tests each day. The positive rate of nucleic acid detection of Covid-19 was determined to be about 10-17%.

**Figure 4:**
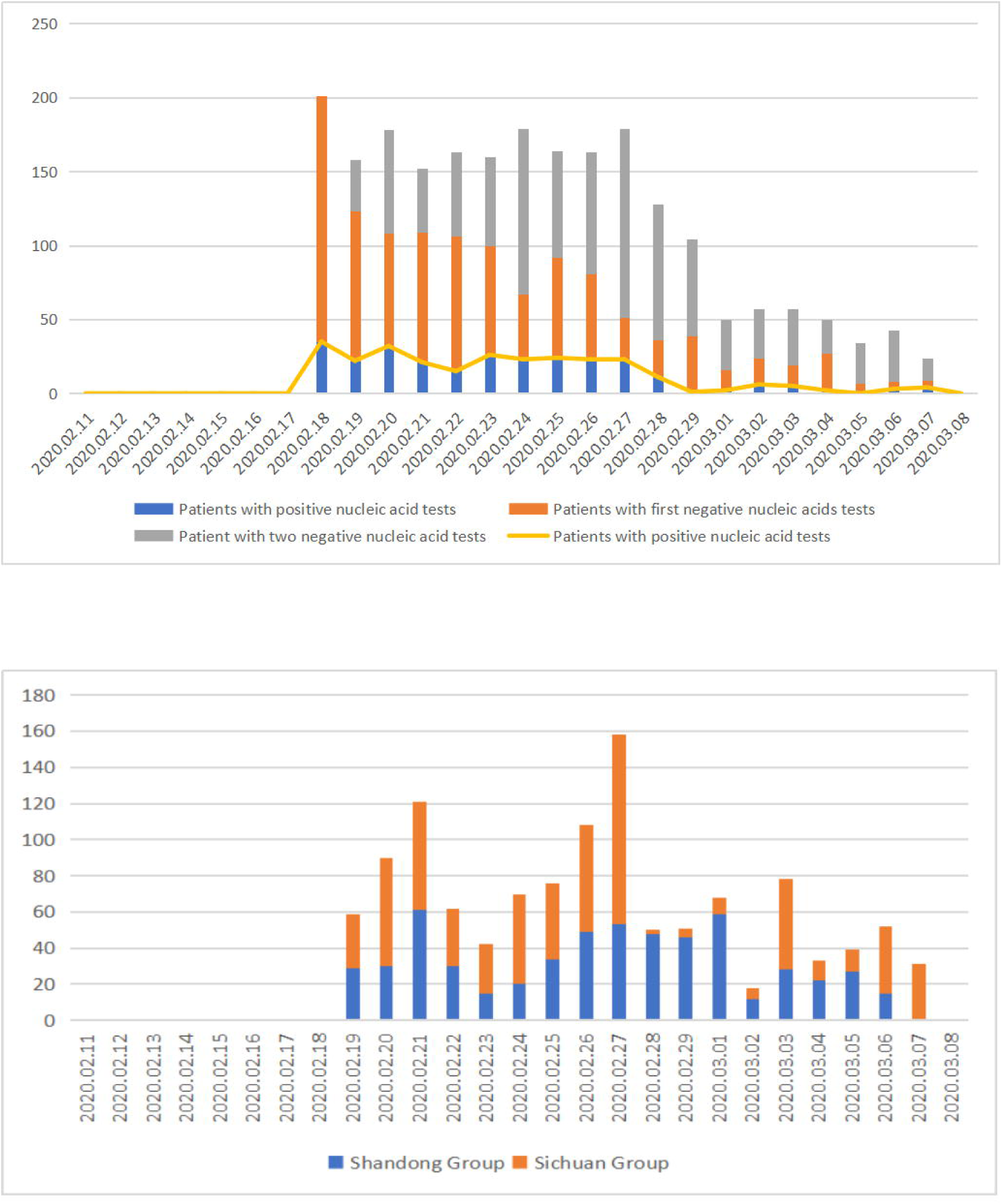
Graph showing the number of nucleic acid detections and CT scan of Covid-19 performed every day. **Figure4a Graph showing the number of nucleic acid detections**. Novel coronavirus nucleic acid testing was performed in the mobile cabin hospital starting on Feb 17th based on the diagnosis and treatment protocol of novel coronaviral pneumonia (trial version 7) in China, when the patients gradually recovered. In this graph, the display of the data of total test results as well as patients showing a negative first nucleic acid test, two negative nucleic acid tests and a positive nucleic acid test. The results indicated there were about 30-120 patients who met discharge criteria with two negative nucleic acid tests each day. The positive rate of nucleic acid detection of Covid-19 was determined to be about 10-17%. Figure 4b: **Graph displaying the number of chest CT scans performed every day** Mobile CT (Shanghai United Imaging HealthCare Co.ltd) were installed and put into use on Feb 19th,8 days after the opening of Hangyang Mobile Cabin Hospital. It indicated the trend for daily CT scans increased every week according to requirements of evaluation of clinical effects which showed the same in total test as well as those in Shandong group (A group) and Sichuan group (B group), the maximum number of CT detection was about 160 times per day.

Mobile CT (Shanghai United Imaging HealthCare Co.ltd) were installed and put into use on Feb 19th,8 days after the opening of Hangyang Mobile Cabin Hospital. It indicated the trend for daily CT scans increased every week according to requirements of evaluation of clinical effects, there were on average about 60-120 CT scans performed, with the maximum number of CT scans being at about 160 times per day for the initial stage, while the data drop to 20-80 due to the decrease of patients.

Over time, there were about 50 patients discharged every day around 10 days later when the patients gradually recovered. Since the newly diagnosed patients dropped dramatically indicating the epidemic was controlled after one incubation period of 14 days (Figure1). No more patients were admitted to the Han Yang MCH from that time onwards.

In total, about 40% of patients were transferred to designated hospitals for treatment due to the severity of Covid-19 infections or other underlying diseases. There were nearly 200 critically ill patients transferred to designated hospitals on March 8, when the cabin hospital was closed because the epidemic in Wuhan had been curbed.

#### IV. Organizational structure of mobile cabin hospitals

Like other mobile cabin hospital, Hayang Mobile Cabin Hospitals had a complete organizational structure to ensure their operation. The head of the local hospital Hubei Women and Children Hospital served as the hospital director for the overall management, the regional government served as district general commander, while the leaders of the different medical teams including Fujian, Sichuan and Shandong Province served as deputy hospital directors to be responsible for supplementary examination, hospital infection control and medical administration, respectively. In addition, the head of another local designated hospital served as the deputy hospital director, thereby ensuring that critically ill patients had a transfer pathway. When in operation, a general commander, medical teams, nursing teams, nosocomial infection teams, and publicity teams were appointed for the mobile cabin hospitals, each of which took charge of work in different domains.

#### V. No nosocomial infections occurred in Hanyang Mobile Cabin Hospital

There were some measures that ensured that no nosocomial infections occurred in Hanyang Mobile Cabin Hospital. First, the division of the “three zones and two passages” was executed appropriately as described above. while as in most MCH, a correct airflow direction could not be made from positive pressure in the clean zone to negative pressure in the contaminated zone in Hanyang MCH in an emergency situation. Secondly, special nosocomial infection teams were formed, consisting of nosocomial infection specialists and part of the nursing staff. 24-hour supervision were performed by this special team for medical staff and other types of staff (e.g. security and cleaning staff) over the proper wearing and removal of protective equipment, to ensure that the prevention and control of nosocomial infections were in place. Thirdly, no material was permitted to be taken out of the contaminated zone in order to ensure no nosocomial infection may occur. Therefore, the data was taken in form of pictures and transferred to headquarters outside the cabin hospital. Lastly but also importantly, medical staff did not go home but returned to their designated hotels after work ensuring that they could not spreading the virus to their family members. Furthermore, medical staff did not take in any food and drinks without doffing PPE when they were working continuously for 6 hours in every shift, once they went back to their residency after work, they conducted immediate personal hygiene, including bathing, eye, ear, mouth and washing the nose, before eating. and timely disinfection of clothing (such as boiling water, or chlorine-containing disinfectant treatment) were performed in order to ensure contamination and infection should not occur in residency.

#### VI. The patients’ livelihoods was safeguarded in mobile cabin hospitals

Mobile cabin hospitals must safeguard the daily lives of all patients, and guaranteeing the livelihoods of nearly a thousand patients in Hanyang mobile cabin Hospital,which was not an easy task. The normal operation of mobile cabin hospitals involved full preparations in terms of livelihood, security guarantee, hygiene, activities and entertainment, and other aspects. Firstly, during the hospitalization and quarantine period, the patients’livehood including clothing, three meals a day, bed linens, paper towels, and other daily necessities, as well as heating systems for the winter were fully managed by the government freely. Secondly, safety protection measures were implemented appropriately. In addition to taking precautions for fire safety, it was also necessary to ensure that conflicts and riots did not occur among patients during hospitalization. These were guaranteed by having a certain ratio of security personnel to patents and staffs. Furthermore, in order to provide patients with an appropriate amount of space for activities, it was also necessary to set up reading rooms, entertainment areas and other facilities to assist in the patients’ daily recreation, thus ensuring their emotional well-being.

During the management of the patients, we adopted a volunteer system, whereby responsible, caring people and activists were selected from each medical unit to serve as unit leaders to assist in management. This enhanced the patients’ daily life management efficiency and orderliness, while also greatly reducing the pressure on medical staff.

#### VII. Treatment regime of patients in mobile cabin hospitals

The adopted main treatment approaches were a combination of traditional Chinese and Western medicine. However, given the limited supply of drugs, the preferred medications were based on traditional Chinese medicine, and the easily available arbidol, and drugs with fewer side effects etc. Biochemical and other tests were not carried out in the mobile cabin hospitals, and hence the side effects of the drugs could not be fully monitored, which was one of the limitations of mobile cabin hospitals. The special treatment approaches adopted in mobile cabin hospitals included psychological treatment, such as providing comfort, alleviation, and social activities(see supplemetal material4). These measures could all effectively reduce patients’ mental anxiety, while also enhancing the enthusiasm of doctors and patients in working harmoniously together to combat the disease.

Based on the discharge criteria in Diagnosis and Treatment Protocol for Novel Coronavirus Pneumonia (Trial Version 7).^[19]^ in China, patients who improved significantly underwent nucleic acid testing(Figure4b) and CT scan(Figure4b). Patients who met the discharged criteria ^[19]^ were discharged, i.e. (1) normal body temperature for more than three days. (2) respiratory symptoms improved significantly. (3) pulmonary imaging showing significant decline of inflammation. (4) negative nucleic acid test result for respiratory pathogens twice consecutively (sampling interval at least one day). Prior to being discharged, the patients’ clothing and luggage were disinfected multiple times. Patients continued to be quarantined at isolation point before going home for another 14 days after discharge to ensure their eventual recovery and minimize the risk of transmission.

### Practical recommendations

The mobile cabin hospitals have the advantage of large capacity, low cost and fast construction speed. Treatment in the cabin hospital mainly included basic medical care and observation of clinical changes, so that severe patients can be transferred to designated hospitals as soon as possible for treatment so as to improve the cure rate ^[20]^. Therefore, they are suitable for an emergency situation in which large-scale spreading occurs at the outbreak of an epidemic when conventional medical resources are not so sufficient. However, if the disease breaks out nationwide and the population has reached a state referred to as herd immunity, the need for cabin hospitals would be greatly reduced ^[21]^. The patients adopted in mobile cabin hospitals included those presenting mild symptoms, with few severe chronic diseases, self-care ability, no mental abnormalities, and active cooperation with medical personnel. In order to ensure the normal operation of the cabin hospital, adequate medical staff, reasonable medical management system and adequate supplies of livehoods were required. Furthermore, the basic mental needs and privacy of patients, and hospital security measures should also be taken into account.

## Discussion

Throughout human history, infectious diseases posed a huge threat to mankind, killing millions of people. This was the case in the 1918 Pandemic, the Black Death, SARS and many others, which killed 50 million, 25 million and 744 people worldwide, respectively ^[22-24]^. In the course of history, there had been precedents using temporary hospitals to fight against epidemics. For example, during the plague outbreak in the Qin Dynasty in China, railway carriages were employed to isolate patients, so as to cut off the source of infection, resulting in curbing the spread of diseases effectively. Moreover, a special infectious hospital (Xiao tang shan Hospital) was established in combating SARS in 2003. Nevertheless, there has no any application of the cabin hospital in dealing with an emergency situation of infectious diseases. COVID-19 was shown to be more highly infectious than any other previous contagious disease ^[25-26]^. In this situation, there had been over ten thousand new cases in less than one month, causing an emergency situation because medical resources could not be increased accordingly. Home quarantine was adopted extensively at an early stage in China for mild patients^[27-28]^ but unfortunately turned out to be ineffective due to the consistent spread of the virus in families in Wuhan. On these basis, in order to cut off the source of infection thoroughly, China initiated to employ cabin hospitals accompanied with a free treatment strategy to achieved the aim of accepting all infected patients, which was shown to be extremely effective, leading to a curbing of the epidemic in Wuhan in only two incubation period of 14 days. There were also other alternative measures such as the use of hotels to provide basic home care for patients with mild cases in Korea, which might have provided a more comfortable livelihood and privacy for patients. Otherwise, this kind of management could only have worked in a setting with limited spreading of the virus but would be difficult to cope with an epidemic which had caused large scale impact. Therefore, in the case of the widespread epidemic the employment of cabin hospital maybe an alternative effective measure to control the infectious diseases.

There were several benefits from the employment of Mobile Cabin Hospitals. With this kind of temporary hospital, all confirmed mild cases received compulsory quarantine in that the sources of infection were cut off quickly and thoroughly when compared to uncontained home quarantine. The strict isolation of patients with mild disease symptoms from the outside world in mobile cabin hospitals helped to reduce the risk of infection among the healthy population and increased the efficiency of centralized diagnosis and treatment by medical staff. As for the patients themselves, this approach reduced the frequency of traveling to and from the hospital, while also preventing transmission to their family members, which slowed down an increase in new cases. Wuhan witnessed this significant effect in only two incubation periods as the newly diagnosed case dropped from over 3 thousand to single digits. Secondly, on one hand, the mild patients were treated at a centralized location and as they were taken care under regular procedures in the Mobile Cabin Hospital, they recovered more rapidly. On the other hand, the severe cases were identified at an early stage and transfer to designated hospital for further treatment, which enabled so many severe cases to be treated effectively as early as possible. Thirdly, relying on the high capacities of mobile cabin hospital for mild cases, sufficient beds in designated hospitals were available for critically ill patients. In addition, following centralized treatment led to a significant increase in recovery rate, a dramatic decrease in mortality rate, and thereby a gradual increase in the number of cured and discharged patients. In summary, by employing cabin hospital, thousands of severe patients were rescued promptly, the cure rate of severe case increased from 14% to 89%. As a result, after 2 incubation periods of using Mobile Cabin Hospital to combat COVID19, the epidemic was effectively curbed, the number of cumulative cases dropped from its peak of more than 80,000 to 14,000. So, all the Cabin Hospital completed their tasks and were closed before Mar 10^[29]^.

In China, 34000 medical staff who aided in Hubei achieved not to get infected due to adhering to the following points. Firstly, everyone was very cautious and attached great importance to it in spirit, so they pursued the personal protection as rigorous as possible. Secondly, China took a very strict attitude to prevent the spread of the virus. Due to the risk of aerosol transmission of the new coronavirus disease^[30]^, all possible transmission routes were cut off, including different entry and exit routes for medical staff and patients, three zones and two channels constructed for prevention of cross infection, doffing PPE layer by layer plus supervision by special team for reducing the risk of self-contamination by PPE, including that all possible contaminated media were being isolated, e.g. all items were not permitted to be taken out, strict three-level protection medical personnel, timely cleaning after work and other measures to ensure that personnel would not be infected. These measures may be regarded as excessive protection, but because of strict adherence, Chinese medical workers were protected well with no cross infection rather than the highly infected rate of about 16% in the United States ^[31]^. Therefore, these preventive measures were worth learning for perfection in the global medical system for infectious diseases.

Although MCH had played an important role in combatting COVID19 in China, in our view, there were some suggestions to improve the feasibility for implementation of MCH:

1. A mature and rapid response management system for infectious disease should be established. Although a powerful management team of mobile cabin hospital was organized by the emergency medical rescue team. The working systems made by the emergency medical team in cooperation with local medical staff and medical staff from nationwide had many defects: they were crude due to temporary thought, many strategies were constantly honed due to immaturity, which consumed much time. If a mature and rapid response management system for infectious disease can be established, a special management team should be trained based on previous experiences, the management efficiency could be greatly improved, medical staff and materials might be greatly reduced too.
2. Some management strategies should be optimized: (1) the structure of the medical staff can be adjusted. Since most patients in MCH were mild patients who could take care of themselves, the core problems of MCH operation were quarantine, basic medical care, arrangement of life and daily management of the patients. The demand of medical staff was not necessarily high. For example, the number of senior professional clinicians could be reduced appropriately, and the number of junior doctors of healthcare staff could be increased to do general care observation, which can greatly reduce the demand for medical personnel (2). Leader patients chosen from every unit to help manage the life of the patients was a useful and effective method to improve management efficiency of MCH and may also reduce the cost and human resources. (3) Intelligent management links were recommended if possible. Automatic simple COVID19 electronic case history system can be filled in before admission to enhance the working efficiency, and the increased use of video instruments in the ward round was able to greatly reduce the exposure of medical staff as well as the use of PPE, and an intelligent program for a screening procedure for nucleic acid detection and CT imaging result could be implemented so as to greatly improve the working efficiency and reduce the number of working staff.
3. It was suggested to establish an emergency management system of the Mobile Cabin Hospital, which was intended help to cut off the source of infection and prevent the further spread of infectious diseases in the early stage of community transmission. The establishment of the emergency management system of mobile cabin hospitals was able to help to deal with large-scale infectious diseases. Different countries may set up emergency management teams for infectious diseases in different regions, and set up emergency response groups for infectious diseases in hospitals at different levels. In this way, the highest administrative unit can mobilize all the medical staff of the emergency teams of hospitals to participate in early stage of epidemic. What is more, sufficient medical personnel can be quickly mobilized to participate in the construction and management of an MCH. Although many countries appeared to have a well-established medical system, they had not prepared for prevention and control of the epidemic of COVID 19, maybe due to not experiencing large-scale infectious diseases in the last 50 years, except countries in Southeast Asia, which were affected by SARS in 2003 and the Middle East by MERS. As a result, the global prevention and control measures of the new epidemic were not effective, causing the continuous spread of the disease. Therefore, it is suggested that the international infectious disease prevention and control system should be strengthened.

After nearly 2 incubation periods of intense efforts, as the number of newly diagnosed cases in Wuhan, the center of epidemic, dropped, the number of new cases in Hubei and China as a whole finally decreased to single digits. Fighting against COVID with mobile cabin hospital was a useful initiative in China and was shown to be effective in handling the COVID19 epidemic ^[20]^, and these experiences may serve as a helpful reference for other countries with epidemic outbreaks.

However, Mobile cabin hospitals also have several notable limitations. For example, they had modest facilities, poor conditions for medical treatment, and a low level of nosocomial infection protection. Therefore, they were not capable of handling severe cases, and it was recommended that only mild cases who meet the admission criteria were accepted. Patients with worsening conditions or underlying diseases that were more challenging to treat should be transferred to designated hospitals for treatment. therefore, once the epidemic had eased, mobile cabin hospitals should be closed as soon as possible, and the main strategy should be switched to admitting and treating patients in designated hospitals ^[32]^.

Currently, the COVID-19 epidemic has spread throughout the world. On March 11, the World Health Organization Director-General Tedros announced that it constituted a global pandemic. At this time, China had controlled the most of the native epidemic for a longer period of time but it also encountered mostly imported case for quite long periods. Therefore, many medical staff members have been working in the front line to prevent the second outbreak of COVID until June 26. As of June 26, a total of 9638755 cases were confirmed worldwide outside of China, with more than 190371 newly diagnosed cases and 5000 new deaths in a single day, as well as a total of more than 487426 deaths ^[32-34]^. We are now living in a time when Covid-19 and other infectious diseases are raging throughout the world. As a community joined by human destiny, the whole world must face this together and jointly launch emergency response strategies. It is our hope that China’s experiences in operating mobile cabin hospitals will help the global response to COVID-19 and other infectious diseases in the future, thus ensuring that the epidemic will come under control as soon as possible.

## Authors Contributors

Conceptualization: Xin Qian, Xinghai Yang.

Data curation: Hongru Li

Formal analysis: Hongru Li

Investigation: Hongru Li, Jiaping Lin, Li Zheng, Xueying Yu, Han Zi, Zhisheng Lin

Methodology: Hongru Li, Li Zheng, Xueying Yu, Han Zi, Zhisheng Lin Lili Ren Project administration: Hongru Li, Xin Qian, Hongmei Lian, Chenkang, Lvyongtao

Writing–original draft: Hongru Li

Writing-review and editing: Hongru Li, Xin Qian, Y Xinghai Yang, Jiaping Lin, Yusheng Chen, Christopher Rensing, Hend A. Wathnani

## Data Availability

All data referred to in the manuscript is available.

## Acknowledgments

This study was supported by the National Major Science and Technology Project for the Control and Prevention of Major Infectious Diseases of China (NO.2017Z×10103004), Fujian Natural Science Foundation (2019J01178), and high-level hospital foster grants from Fujian Provincial Hospital, Fujian province, China (NO.2019HSJJ11). We also acknowledge Researcher Supporting Project number (RSP-2020/205), King Saud University, Riyadh, Saudi Arabia. We would like to thank Editage (www.editage.cn) for English language editing.

